# Field comparison of STANDARD™ Q Filariasis Antigen Test (QFAT) with Bioline Filariasis Test Strip (FTS) for the detection of Lymphatic Filariasis in Samoa, 2023

**DOI:** 10.1101/2024.04.01.24305170

**Authors:** Jessica L Scott, Helen J Mayfield, Jane Sinclair, Beatris Mario Martin, Maddison Howlett, Ramona Muttucumaru, Kimberly Y Won, Robert Thomsen, Satupaitea Viali, Rossana Tofaeono-Pifeleti, Patricia M Graves, Colleen L Lau

**Author notes:** **Corresponding author** (JLS), (CLL).

## Abstract

**Background:** To monitor the progress of lymphatic filariasis (LF) elimination programmes, field surveys to assess filarial antigen (Ag) prevalence require access to reliable, user-friendly rapid diagnostic tests. We aimed to evaluate the performance of the new Q Filariasis Antigen Test (QFAT) with the currently recommended Filariasis Test Strip (FTS) for detecting the Ag of *Wuchereria bancrofti*, the causative agent of LF, under field conditions.

**Methodology/Principal Findings:** During an LF survey in Samoa, 344 finger-prick blood samples were tested using FTS and QFAT. Microfilariae (Mf) status was determined from blood slides prepared from Ag-positive samples. Each test was re-read at 1 hour and the next day to determine the stability of results over time. Overall Ag-positivity by FTS was 29% and 30% by QFAT. Concordance between the two tests was 94% (Kappa=0.85). Of the 105 Mf slides available, 38.1% were Mf-positive, and all were Ag-positive by both tests. Darker test line intensities from Ag-positive FTS were found to predict Mf-positivity (compared to same/lighter line intensities). QFAT had significantly higher reported test result changes than FTS, mostly reported the next day. The field laboratory team preferred QFAT over FTS due to the smaller blood volume required, better usability, and easier readability.

**Conclusion/Significance:** QFAT could be a suitable and user-friendly diagnostic alternative for use in the monitoring and surveillance of LF in field surveys based on its similar performance to FTS under field conditions.

**Author Summary:** Lymphatic filariasis (LF) is a debilitating tropical disease caused by an infection with parasitic filarial worms that are transmitted by mosquitoes. Long-term infection can lead to stigmatising chronic conditions like lymphoedema and elephantiasis. The World Health Organization initiated the global programme to eliminate lymphatic filariasis (GPELF) in 2000, which focuses on the mass administration of anti-LF drugs to stop transmission in endemic countries. However, to monitor the success of this programme and to make informed decisions to stop costly mass drug administrations, it is crucial to have access to accurate and reliable rapid diagnostics. Here, we evaluated the performance of a new rapid antigen test called the Q Filariasis antigen test and compared it to the currently recommended filariasis test strip under field conditions in Samoa. This study showed that the new rapid test could be a suitable alternative to the currently recommended test for use in GPELF-related activities with more user-friendly features.

## Introduction

Lymphatic filariasis (LF) is a mosquito-borne neglected tropical disease caused by infection with a parasitic worm. Consequences of long-term infection include chronic disabling and disfiguring manifestations such as lymphoedema, including scrotal hydrocele, and elephantiasis. These morbidities could lead to social stigmatisation and loss of work (1). The main pathogen causing LF is the filarial worm *Wuchereria bancrofti* and, to a lesser extent, *Brugia malayi* and *B. timori* (2).

To eliminate LF as a public health problem, the World Health Organization (WHO) established the Global Programme to Eliminate Lymphatic Filariasis (GPELF) in 2000. One arm of the program is focused on interrupting the transmission of LF through repeated rounds of mass drug administration (MDA) of anti-filarial drugs in endemic regions (3). In conjunction with the GPELF, the Pacific Programme to Eliminate LF (PacELF) was launched to support the 16 endemic Pacific Island Countries and Territories committed to combating this disease (4). Since 1999, the elimination of LF as a public health problem has been validated in eight countries in the PacELF region, while the other eight, including Samoa, remain committed to eliminating the disease. Accessibility to robust and reliable diagnostic tools remains essential for monitoring the progress of GPELF activities and for post-validation surveillance (5).

In regions where *W. bancrofti* is the leading causative agent of LF, WHO recommends the qualitative Alere Bioline Filariasis Test Strip (FTS; Alere Abbott), a rapid diagnostic test for detecting circulating filarial antigen in human blood samples. Since 2015, FTS has been successfully used in national programs, replacing the former immunochromatographic test (ICT, BinaxNOW). Previous studies reported that FTS was preferred over ICT, as it was more stable in the field, cheaper, and able to detect lower concentrations of circulating filarial antigen than ICT (6), but its useability and user-friendliness under field conditions were considered a drawback as it is susceptible to user-error (6, 7). Later, concerns were raised regarding potential cross-reactivity with other filarial species, such as *Loa loa* (8), although this parasite is not endemic in the Asia-Pacific. Further, since the COVID-19 pandemic, relying on a single manufacturer has introduced procurement challenges for rapid diagnostic tests, which has been a barrier for LF programmatic surveys.

A new rapid antigen test, the Q Filariasis Antigen test (QFAT) (SD Biosensor, Suwon, South Korea), has been proposed as an alternative to the FTS for GPELF-related LF surveys. This new test also detects filarial antigens from capillary blood, like FTS and requires less sample volume, but its utility under field conditions has not been evaluated. This study aimed to evaluate the performance of the new QFAT and FTS when deployed in an endemic region under field conditions. The specific objectives were to compare the concordance between the two tests for detecting the circulating antigen of *W. bancrofti*, to assess whether the intensity of the test line of QFAT could indicate microfilariae positivity and determine test stability over time by re-reading the tests at one hour and the next day. The findings from this study will support recommendations regarding the suitability of QFAT as an alternative field diagnostic for GPELF-related activities.

## Materials and methods

### Study setting and participants

Field and laboratory work for this study was undertaken in Samoa, on the main island of Upolu, in 2023. Samples used in this study were sourced from three study components associated with a monitoring and surveillance field project for LF. The first study component included follow-up of participants (and their household members) from surveys conducted in Samoa in 2018 and 2019 (9, 10), who tested antigen (Ag) positive and had detectable microfilaria (Mf) in blood, as identified by microscopy. The second component was a community-based survey of eight sentinel villages, which were selected based on Ag prevalence in 2019 (two villages, each with zero, low (3-4%), medium (6-7%) and high (13-16%) prevalence). The final component involved targeted testing of households neighbouring those Ag-positive participants identified in the 2019 survey.

### Blood sample collection and processing

Heparinised microvettes were used to collect 300μL of capillary (finger-prick) blood samples from participants. Immediately after collection, blood samples were stored in a cold storage container until the samples were delivered to the field laboratory, where the samples were stored in the fridge at 4°C. Typically, samples were processed and tested the next morning.

Before testing, samples were acclimated to room temperature and tested in a field laboratory. The rapid tests were performed per the manufacturer’s instructions using 75μL of blood for FTS and 20μL for QFAT. The precise volumes were transferred to the respective tests using a calibrated micropipette.

### Reading and interpretation of rapid antigen test results

Results of FTS and QFAT were read at the manufactures recommended time of 10 minutes by up to three independent readers. Occasionally, a high laboratory workload meant that it was not possible for all tests to be read by three readers. Tests were read by the naked eye and illuminated with a torch if needed. Results were classified by each reader as positive, negative, or invalid (no sample flow or absence of control line). Samples with invalid results were repeated if there was sufficient blood. If the second test produced a valid result, this was recorded as the final result. If the second test produced another invalid result, it was recorded as invalid.

At the 10-minute readings, tests with a positive result had the test line semi-quantified based on the intensity of the test line compared to the control line. These tests were scored by up to three readers, accordingly: (1) test lines were lighter than the control line; (2) test lines were the same intensity as the control line; or (3) test lines were darker than the control line. Each test was then re-read at 1 hour and the next day (e.g. 12-18 hours) by one reader to determine whether the tests remained stable over time. Re-reading the Ag tests after 10 minutes is not recommended by the manufacturers.

### Selection of blood samples for QFAT comparison

To select the samples for the QFAT trial, all samples from the wider survey were tested with FTS in batches of five. Any batch of five samples that included at least one FTS-positive sample was also tested using QFAT (Figure 1). This strategy ensured sufficient numbers of Ag-positive samples for comparisons between FTS and QFAT. Therefore, it is important to note that the Ag positivity rate presented in this study does not represent the prevalence in any of the three study components mentioned previously. In addition to the above selection strategy, 14 samples that were inadvertently frozen and invalid by FTS were purposefully included in the QFAT trial for comparison.

**Figure 1:**
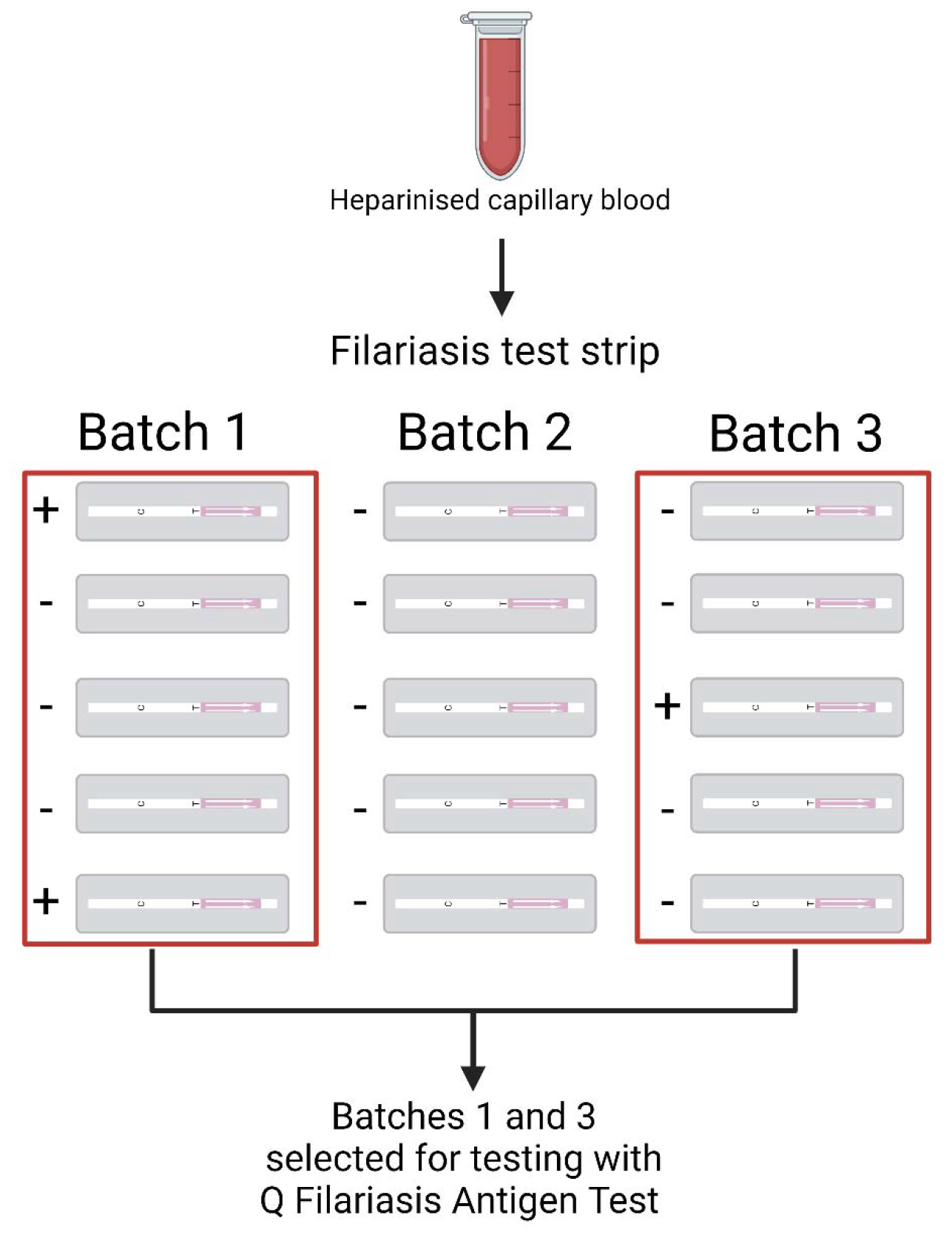
Strategy used to select samples for Q Filariasis antigen test (QFAT) evaluation. Batches of Filariasis test strip with at least one antigen-positive test were selected for testing using QFAT. ‘+’ indicates antigen (Ag)-positive tests and ‘-’ indicates Ag-negative tests. Image created with BioRender.com.

### Determination of Mf status from thick smear blood films

Blood samples testing positive for LF Ag by FTS and/or QFAT, up to three thick smear slides were prepared (depending on the blood volume available). Each slide included three 20 μL stripes of blood. Slides were dried, dehaemoglobinised in water and stained with Giemsa following standard procedures (11). Two independent blinded readers examined at least two slides per participant. If at least one slide reader detected any Mf, it was classified as Mf-positive. If neither slide reader detected Mf, it was classified as Mf-negative. Slides with discordant Mf results between the two slide readers were resolved by jointly re-examining the slides and making a final judgment.

### Data analysis

Data was analysed using GraphPad Prism 9 (Prism for Windows, version 9.2.0.332). FTS and QFAT summaries were described as frequencies and percentages and reported with a 95% confidence interval (CI). Final interpretations of the test results for FTS and QFAT were determined by the consensus between the readers and were done accordingly; tests with discordant readings between readers, the dominant reading was used as the result. If the readings were discordant for tests with only two readers, it was classified as indeterminant. In cases where a single reader provided the interpretation, this reading was considered final. Discordance between the readers interpretations was determined when test interpretations made by the readers differed from one another and the analysis excluded samples with invalid interpretations.

Concordance between the results of the two rapid Ag tests (agreement or disagreement between positive and negative results) was determined and reported as a percentage. Cohen’s Kappa (K) agreement statistics were performed to determine the probability that agreement between the tests was not due to chance and reported with a 95% CI. Line intensity scores from Ag-positive tests to use for the analysis were selected based on the consensus of test line intensity scores by the independent readers. If no consensus was reached by the readers for a particular test result, the result was excluded. These line intensity scores for FTS and QFAT were then compared for those samples where Mf-status was determined. A univariable logistics regression analysis was conducted to assess whether the darker test lines of Ag-positive tests (compared to those with equal or lesser intensity than the control line) were predictive of Mf-positivity. The relationship was presented as an odds ratio (OR) and reported with a 95% CI.

FTS and QFAT tests were re-read at 1 hour and next day time points to determine whether the test results changed over time (from Ag-negative to Ag-positive, or vice versa). Tests with valid positive or negative results at all three time points were compared. The McNemar test was used to determine the difference between the proportion of tests that changed results from 10 minutes to 1 hour, and from 1 hour to the next day. Fisher’s exact test was used to determine the difference between the proportion of tests that changed results for FTS and QFAT at 1 hour and next day readings. All statistical inferences were based on a p-value of <0.05.

### QFAT usability under field conditions

Four field laboratory workers, trained before sample testing, provided independent verbal feedback on FTS and QFAT at the conclusion of the field comparison. Two of the four laboratory workers have had previous experience using FTS and/or QFAT. Laboratory workers were asked for feedback on test set-up, sample application and volume, test readability and other relevant aspects of the test’s characteristics and procedure.

## Results

A total of 344 whole blood samples were tested using both FTS and QFAT and were used to evaluate test concordance at the initial reading (10 minutes after application of the sample). At the 10-minute reading, no indeterminant results were reported for QFAT, but three (1.0%) were reported for FTS. Of the 14 (4.1%) samples that were invalid by FTS, none were reported as invalid by QFAT. For the invalid FTS readings, no control lines were reported for the tests, even after repeating the samples on a newer batch of test strips. All 14 samples invalid by FTS produced a valid result by QFAT (one positive and thirteen negatives). Thick blood smears were prepared for 105 Ag-positive participants, and 40 (38.1%, 95% CI 29.4-47.6%) of these were classified as Mf-positive. After the initial reading, 309 (89.8%) of the FTS and 341 (99.1%) of QFAT were read again at 1 hour and the next day and were used for further analysis.

### Number of readers and discordance between readers at initial reading

The number of readers for FTS and QFAT at the initial reading (10 minutes) and discordance between the readers is summarised in Table 1. Among the 344 samples evaluated and removal of the 14 invalid FTS interpretations, 70.3% of FTS and 84.6% of QFAT were read by three independent blinded readers. The total proportion of interpretation discordance between readers was highest for QFAT (4.4%) than FTS (2.1%). Specifically, for tests with only two readers, there was higher discordance for FTS (1.0%) than QFAT (0%), but for tests with three readers, discordance for QFAT (4.4%) was higher than FTS (1.2%).

**Table 1:**
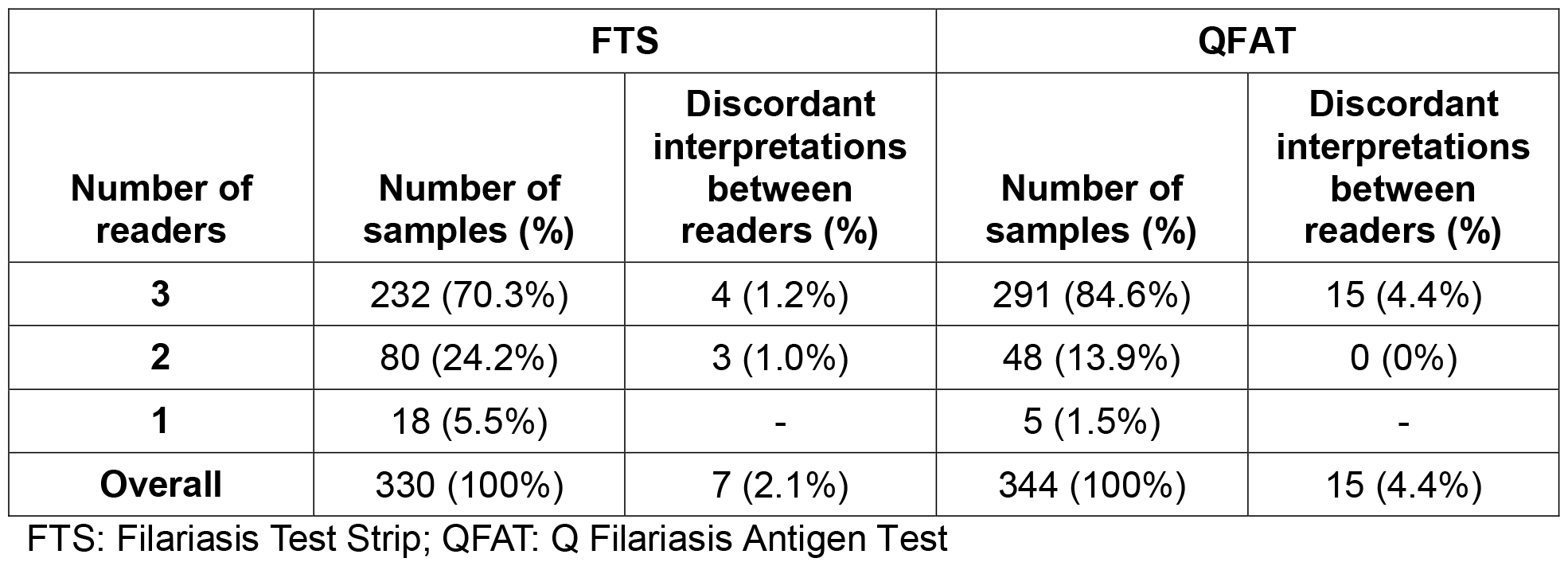
Number of readers for Filariasis Test Strip and Q Filariasis Antigen Test at initial reading (at 10 minutes) and discordant results between readers, excluding invalid test interpretations, Samoa 2023.

|  | FTS |  | QFAT |  |
| --- | --- | --- | --- | --- |
| Number of readers | Number of samples (%) | Discordant interpretations between readers (%) | Number of samples (%) | Discordant interpretations between readers (%) |
| 3 | 232 (70.3%) | 4 (1.2%) | 291 (84.6%) | 15 (4.4%) |
| 2 | 80 (24.2%) | 3 (1.0%) | 48 (13.9%) | 0 (0%) |
| 1 | 18 (5.5%) | - | 5 (1.5%) | - |
| Overall | 330 (100%) | 7 (2.1%) | 344 (100%) | 15 (4.4%) |

FTS: Filariasis Test Strip; QFAT: Q Filariasis Antigen Test

### LF antigen positivity by FTS and QFAT

Overall, 100 samples tested positive with FTS (29.0%, 95% CI 24.5-34.1%) and 104 samples with QFAT (30.2%, 95% CI 25.6-35.3%). If indeterminant and invalid FTS results were excluded from the analysis, Ag positivity was 30.6% (95% CI 25.8-35.8%) by FTS and 30.2% (95% CI 25.6-35.3%) by QFAT.

### FTS and QFAT concordance at initial reading (at 10 minutes)

The overall concordance between FTS and QFAT, including indeterminant and invalid readings (n=344), was 93.6%. The Kappa agreement statistic indicated excellent agreement between the two tests (K=0.85; 95% CI 0.80-0.91). Table 2 demonstrates that of the valid but discordant test results, one was positive by FTS and negative by QFAT, and four were positive by QFAT and negative by FTS.

**Table 2:** Concordance between Filariasis Test Strip and Q Filariasis Antigen Test results, including indeterminant and invalid tests, Samoa 2023.

|  |  | FTS |  |  |  |  | Concordance (%) | Kappa (95% CI) |
| --- | --- | --- | --- | --- | --- | --- | --- | --- |
|  |  | Positive | Negative | Indeterminant | Invalid | Total |  |  |
| QFAT | Positive | 99 | 4 | 0 | 1 | 104 | 93.6 | 0.85 (0.80-0.91) |
|  | Negative | 1 | 223 | 3 | 13 | 240 |  |  |
|  | Indeterminant | 0 | 0 | 0 | 0 | 0 |  |  |
|  | Invalid | 0 | 0 | 0 | 0 | 0 |  |  |
|  | Total | 100 | 227 | 3 | 14 | 344 |  |  |

CI: Confidence interval; FTS: Filariasis Test Strip; QFAT: Q Filariasis Antigen Test

If indeterminant and invalid readings were excluded from the analysis, the concordance between FTS and QFAT (n=327) improved to 98.5% (Table 3). The Kappa agreement similarly indicated excellent agreement between the two tests (K=0.96; 95% CI 0.96-1.0).

**Table 3:**
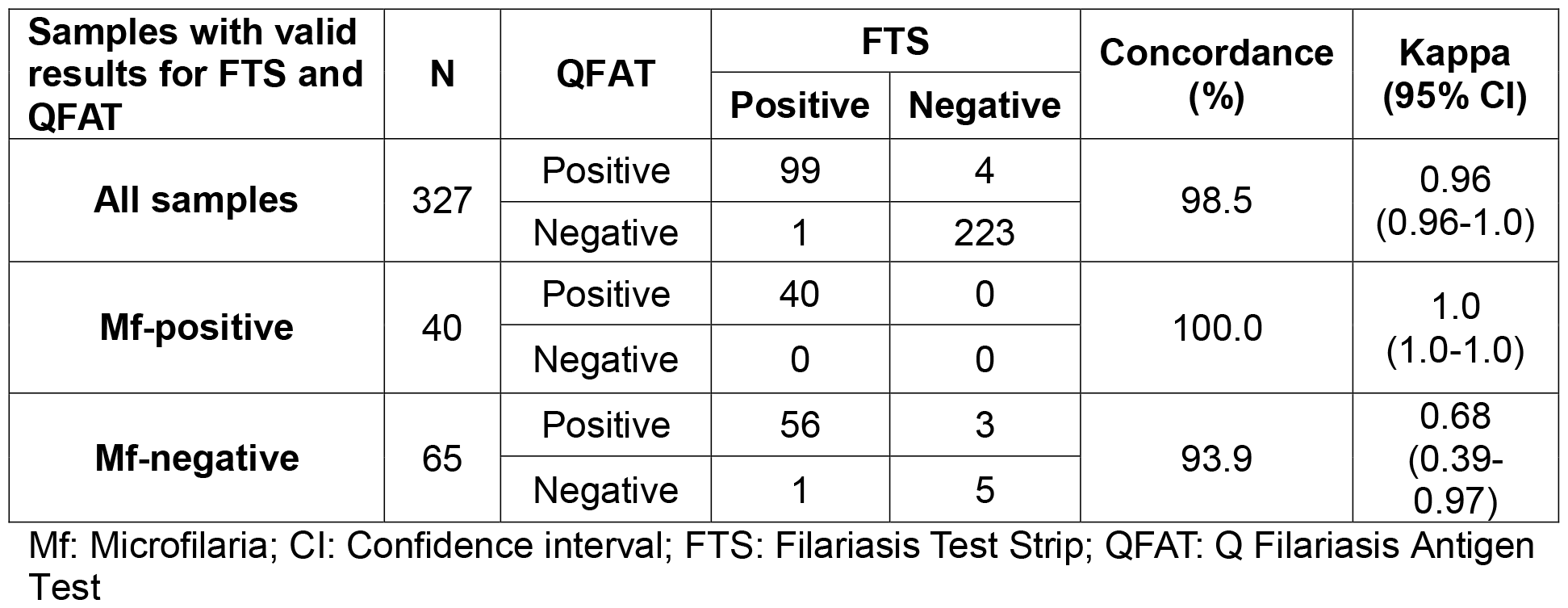
Concordance between Filariasis Test Strip and Q Filariasis Antigen Test for all samples and stratified by microfilaria positive or negative, excluding indeterminant and invalid results, Samoa 2023.

| Samples with valid results for FTS and QFAT | N | QFAT | FTS |  | Concordance (%) | Kappa (95% CI) |
| --- | --- | --- | --- | --- | --- | --- |
|  |  |  | Positive | Negative |  |  |
| All samples | 327 | Positive | 99 | 4 | 98.5 | 0.96 (0.96-1.0) |
|  |  | Negative | 1 | 223 |  |  |
| Mf-positive | 40 | Positive | 40 | 0 | 100.0 | 1.0 (1.0-1.0) |
|  |  | Negative | 0 | 0 |  |  |
| Mf-negative | 65 | Positive | 56 | 3 | 93.9 | 0.68 (0.39-0.97) |
|  |  | Negative | 1 | 5 |  |  |

Mf: Microfilaria; CI: Confidence interval; FTS: Filariasis Test Strip; QFAT: Q Filariasis Antigen Test

All 40 Mf-positive samples were Ag-positive by both FTS and QFAT. Of the 65 Mf-negative samples, 56 (86.1%) were Ag-positive by both QFAT and FTS, with 93.9% concordance and good agreement (K=0.68; 95% CI 0.39-0.97) between the two tests (Table 3).

### Antigen-positive FTS and QFAT test line intensity scores

The distribution of the FTS and QFAT test line intensity scores among the Mf-positive and Mf-negative participants are presented in Figure 2. Blood samples that were Ag-positive by FTS and Mf-positive produced predominantly strong (3) test lines (86.5%), compared to FTS Ag-positive/Mf-negative samples. In contrast, blood samples testing Ag-positive by QFAT and Mf-positive had mostly light (1) intensity test lines (92.4%), similar to test line intensities reported for QFAT Ag-positive and Mf-negative blood samples. For samples that tested positive for Ag by FTS, those with test lines darker than the control had 13.6 times higher odds of being Mf-positive (95% CI 4.8-45.4), compared to those with test lines that were the same or lighter intensity than the control (p-value <0.0001). However, a logistics regression could not be applied for QFAT because there were no samples with test lines that were darker than the control line.

**Figure 1:**
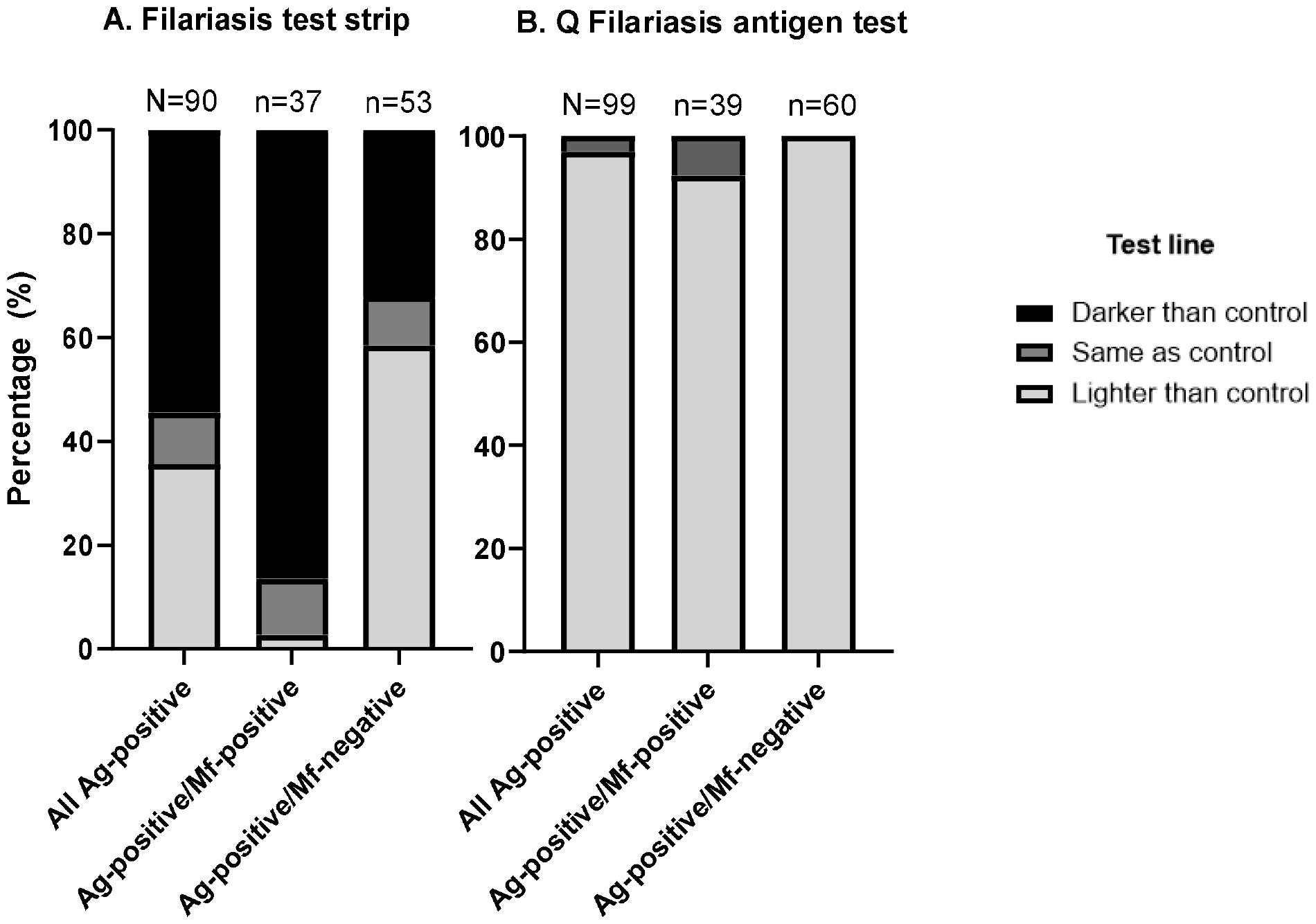
Summary of test line gradings (darker, same or lighter than control line) determined for participants with Antigen (Ag)-positive (A) filariasis test strip and (B) Q Filariasis antigen tests, against microfilariae (Mf) status

### Changes in FTS and QFAT test results at 1 hour and next day timepoints

We assessed the difference between FTS and QFAT test results at three time points (10-minutes, 1 hour and next day) to determine whether test results remained stable over time. Only data sets with complete and valid readings across all three time points were included in the analysis. Ag positivity at 10 minutes was 30.1% by FTS and 29.9% by QFAT (Supplementary 1, Table 4). At 1 hour, Ag positivity was 30.2% by FTS and 29.6% by QFAT. The next day, the Ag positivity remained at 30.2% for FTS but increased to 39.6% for QFAT.

A total of 46 (13.5%) QFAT tests were observed to have results change between 10 minutes and 1 hour and/or the next day, while five (1.6%) changed for FTS. The proportion of QFAT tests that had a reported change of results between the 1 hour and next day readings was statistically significant (McNemar test p-value = 0.003). Additionally, there was a statistically significant difference between the proportion of tests changing for FTS and QFAT at the next-day readings (Fisher’s exact test p-value = 0.006). These results indicate minor changes in test result readings for FTS and QFAT between 10 minutes and 1 hour, whereas, at the next-day readings, the number of test results reported to have changed was significantly greater for QFAT than for FTS. The distinct proportion of tests reported to have changed at these times points for FTS and QFAT have been broken down in supplementary file 2.

### QFAT usability under field conditions

The field laboratory team reported that QFAT was preferred over FTS for multiple reasons. QFAT required smaller sample volumes than FTS. Further, the smaller sample volume required for QFAT made it easier to apply the blood sample, as blood spillage off the application pad was occasionally noted with FTS but not QFAT. The QFAT cassette was easier to handle than the loose strip and tray used for the FTS, and QFAT occupied less space in the field laboratory. Furthermore, the control line for QFAT was consistently clearer and easier to identify than for FTS. Although the test line on QFAT sometimes appeared lighter than the control, it was still identifiable even without a torch, unlike FTS, which could be challenging to see at times. It was also noted that in times of high laboratory demand the additional buffer step required for QFAT could be missed, which could potentially lead to invalid test results.

## Discussion

Our study found excellent concordance in LF Ag results between FTS and QFAT. Ag positivity based on initial 10-minute readings was similar for both tests, with a slightly higher reported positivity rate for QFAT. Additionally, it was encouraging to find that all Mf-positive samples were Ag-positive by both QFAT and FTS as these samples tend to have higher levels of detectable circulating Ag (12). Discordance between the independent readers was highest for QFAT compared to FTS. However, all tests were resolvable due to the consensus among readers, except for 1.0% of FTS test results that were classified as indeterminant.

It has been previously suggested that semi-quantification of the test line as compared to the control line could indicate Mf-positivity from blood samples (13), which could have potential utility in field studies to indirectly infer the level of Mf rates in a population over time (14). In this current study we found that darker test line intensities, compared to the control line, produced by Ag-positive FTS was a predictor of Mf-positivity. However, this was not the case for QFAT, as no samples produced a test line that was darker than the control line. Indicating that the semi-quantification of the test line for Ag-positive QFAT tests may have limited utility for indicating Mf-positivity. However, this finding could be influenced by the smaller sample volume required for QFAT than for FTS, as the greater intensity of the FTS test line noted in this study could potentially be influenced by the tests greater sample volume requirement. It should also be noted that QFAT consistently had stronger control lines as compared to FTS, whereby the control line of FTS was often very faint. The darker controls lines are more reliable as they are less prone to misinterpretations and being discarded as invalid.

A significantly greater proportion of tests were reported to have changed for QFAT than FTS at the next day readings. The propensity of the reported change was negative to positive. A previous study comparing and assessing the stability of the former ICT with FTS over time revealed that participants from a non-endemic country had Ag-negative FTS tests at 10 minutes, but after 24 hours, some of the tests turned positive (6), indicating some level of unreliability of FTS if test results are read after the recommended time. The findings from this study suggest that test results from QFAT could produce false-positives overtime, indicating that results should not be interpreted beyond the manufacturer’s recommended timeframe.

In general, the field laboratory team preferred QFAT over FTS predominantly due to its smaller sample volume requirements, which would be beneficial to field workers as it allows sufficient volumes to be used for other purposes (e.g., Mf slides and/or dried blood spots) or for repeating FTS or QFAT if needed, but also for its user-friendliness. However, the evaluation of user-friendliness may be limited as micropipettes were used to apply precise volumes to the test sample pad, deviating from the capillary tubes provided by the test kits.

The strengths of this study include the ability to simulate the performance of QFAT under field conditions, given its integration within an LF community survey. However, in LF programmes QFAT is intended for point-of-care application, specifically using blood directly from a finger-prick. This approach was not evaluated in this current study. While QFAT has demonstrated promising performance under field conditions in Samoa, additional field evaluations are recommended for other settings and should consider evaluating the point-of-care aspect. Additionally, having multiple readings of the test results at the initial 10 minutes was an advantage to this study. Multiple readers enabled the resolution of discordant results, enhancing the reliability of the conclusions drawn from this study. Cross-reactivity was not explored in this current study and while concerns of cross-reactivity of FTS with *L. loa* (8) and potentially strongyloidiasis (6) have been raised, it is unknown whether this is an issue for QFAT. Other remaining knowledge gaps include whether QFAT will have comparable results to FTS after repeated rounds of MDA and/or where Ag prevalence low. Additionally, a cost-benefit analysis would also be of interest to determine whether QFAT’s accuracy and user-friendliness demonstrated in this study leads to cost savings compared to using FTS.

In summary, QFAT demonstrated promising performance under field conditions in Samoa. Our study found that QFAT and FTS Ag-positivity rates were comparable, with excellent concordance between the two tests. The field laboratory team preferred QFAT over FTS due to smaller sample volume requirements, ease of use, and clearer readability, offering reliable and user-friendly Ag detection test for LF surveillance and control programs.

## Data Availability

All data produced in the present study are available upon reasonable request to the authors. All data produced has also been lodged with the Task Force for Global Health, and summary data will be available on request if approved by the Samoa Ministry of Health.

## Abbreviations

Ag: Antigen
LF: Lymphatic filariasis
QFAT: Q filariasis antigen test
FTS: Filariasis test strip
Mf: Microfilariae
WHO: World Health Organization
GPELF: Global Programme to Eliminate Lymphatic Filariasis
MDA: Mass drug administration
PacELF: Pacific Programme to Eliminate Lymphatic Filariasis
ICT: Immunochromatographic test
CI: Confidence interval
K: Cohen’s Kappa

## Acknowledgements

We are grateful to the all the participants who took part in this survey. We are also deeply appreciative of the Samoa Red Cross team and Stephanie Curtis for their assistance in the conducting the field work, which formed an integral part of this study. We would like to thank the SD biosensor for providing the QFATs and extend our gratitude to Jonathan King, who was the liaison between NTD-WHO, PQ, the procurement departments and the manufacturers.

## Conflicts of Interest

Nothing to declare.

## Funding

This work received financial support from the Coalition for Operational Research on Neglected Tropical Diseases (COR-NTD), which is funded at The Task Force for Global Health primarily by the Bill & Melinda Gates Foundation (OPP1190754), by UK aid from the British government, and by the United States Agency for International Development through its Neglected Tropical Diseases Program. Under the grant conditions of the Foundation, a Creative Commons Attribution 4.0 Generic License has already been assigned to the Author Accepted Manuscript version that might arise from this submission. CLL was supported by an Australian National Health and Medical Research Council Fellowship (APP 1193826).

## Ethical Approval Statement

Ethics approvals were obtained from the Samoa Ministry of Health and the Human Research Ethics Committee (HREC) of The University of Queensland (protocol 2021/HE000895), and the Australian National University HREC provided recognition of approval by another HREC.

## Disclaimer

The findings and conclusions in this report are those of the authors and do not necessarily represent the official position of the Centers for Disease Control and Prevention.

## Declaration of Generative AI and AI-assisted technologies in the writing process

During the preparation of this work the first author used Grammarly® for Microsoft Office (v.6.8.263) to check grammar and spelling. After using this tool, the author reviewed and edited the content as needed and takes full responsibility for the content of the publication.

## Notes

### Competing Interest Statement

The authors have declared no competing interest.

## References

1. Taylor MJ, Hoerauf A, Bockarie M. Lymphatic filariasis and onchocerciasis. The Lancet. 2010;376(9747):1175–85.

2. WHO. Lymphatic filariasis: World Health Organization (WHO); 2023 [Available from: https://www.who.int/news-room/fact-sheets/detail/lymphatic-filariasis.

3. WHO. Ending the neglect to attain the sustainable development goals: a road map for neglected tropical diseases 2021–2030. World Health Organization (WHO); 2020. Report No.: 9240018794.

4. Ichimori K, Graves PM. Overview of PacELF—the Pacific Programme for the Elimination of Lymphatic Filariasis. Trop Med Health. 2017;45(1):34.

5. Rebollo MP, Bockarie MJ. Shrinking the lymphatic filariasis map: update on diagnostic tools for mapping and transmission monitoring. Parasitology. 2014;141(14):1912–7.

6. Weil GJ, Curtis KC, Fakoli L, Fischer K, Gankpala L, Lammie PJ, et al. Laboratory and field evaluation of a new rapid test for detecting Wuchereria bancrofti antigen in human blood. Am J Trop Med Hyg. 2013;89(1):11–5.

7. Pantelias A, King JD, Lammie P, Weil GJ. Development and Introduction of the Filariasis Test Strip: A New Diagnostic Test for the Global Program to Eliminate Lymphatic Filariasis. Am J Trop Med Hyg. 2022;106(5_Suppl):56–60.

8. Hertz MI, Nana-Djeunga H, Kamgno J, Jelil Njouendou A, Chawa Chunda V, Wanji S, et al. Identification and characterization of Loa loa antigens responsible for cross-reactivity with rapid diagnostic tests for lymphatic filariasis. PLoS Negl Trop Dis. 2018;12(11):e0006963.

9. Lau CL, Meder K, Mayfield HJ, Kearns T, McPherson B, Naseri T, et al. Lymphatic filariasis epidemiology in Samoa in 2018: Geographic clustering and higher antigen prevalence in older age groups. PLoS Negl Trop Dis. 2020;14(12):e0008927.

10. Graves PM, Sheridan S, Scott J, Amosa-Lei Sam F, Naseri T, Thomsen R, et al. Triple-Drug Treatment Is Effective for Lymphatic Filariasis Microfilaria Clearance in Samoa. Trop Med Infect Dis. 2021;6(2):44.

11. WHO. Lymphatic filariasis: monitoring and epidemiological assessment of mass drug administration: a manual for national elimination programmes. Geneva, Switzerland: WHO: World Health Organization (WHO); 2011.

12. Chesnais CB, Vlaminck J, Kunyu-Shako B, Pion SD, Awaca-Uvon NP, Weil GJ, et al. Measurement of Circulating Filarial Antigen Levels in Human Blood with a Point-of-Care Test Strip and a Portable Spectrodensitometer. Am J Trop Med Hyg. 2016;94(6):1324–9.

13. Chesnais CB, Missamou F, Pion SDS, Bopda J, Louya F, Majewski AC, et al. Semi-Quantitative Scoring of an Immunochromatographic Test for Circulating Filarial Antigen. Am J Trop Med Hyg. 2013;89(5):916–8.

14. Nana-Djeunga HC, Sicard CM, Mogoung-Wafo AE, Chesnais CB, Deléglise H, Touka-Nounkeu R, et al. Changes in Onchocerciasis Ov16 IgG4 Rapid Diagnostic Test Results Over One-Month Follow-up: Lessons for Reading Timeframe and Decision-Making. Am J Trop Med Hyg. 2022;107(3):658–61.

