## Supplemental Table 1 for "Field comparison of STANDARD™ Q Filariasis Antigen Test (QFAT) with Bioline Filariasis Test Strip (FTS) for the detection of Lymphatic Filariasis in Samoa, 2023"

Supplementary 1

Table 4: Antigen positivity at the time intervals, 10 minutes, 1 hour and next day. Total sample size included for this analysis were 300 for FTS and 341 for QFAT.

| **Timepoints** | **FTS** | | **QFAT** | |
| --- | --- | --- | --- | --- |
|  | **Number of tests positive** | **% (95% CI)** | **Number of tests positive** | **% (95% CI)** |
| **10 minutes** | 93 | 30.1 (25.3-35.4) | 102 | 29.9 (25.3-35.0) |
| **1 hour** | 94 | 30.2 (25.4-35.5) | 101 | 29.6 (25.0-34.7) |
| **Next day** | 94 | 30.2 (25.4-35.5) | 135 | 39.6 (34.5-44.9) |

CI: 95% confidence interval; FTS: Filariasis Test Strip; QFAT: Q Filariasis Antigen Test
