## Supplemental Figure 1 for "Field comparison of STANDARD™ Q Filariasis Antigen Test (QFAT) with Bioline Filariasis Test Strip (FTS) for the detection of Lymphatic Filariasis in Samoa, 2023"

**Supplementary 2**

Figure 3: Changes in readings for FTS (A) and QFAT (B) from the initial 10-minute reading to 1 hour and next-day readings post application of blood.

**10 minutes**

**1 hour**

**Next day**

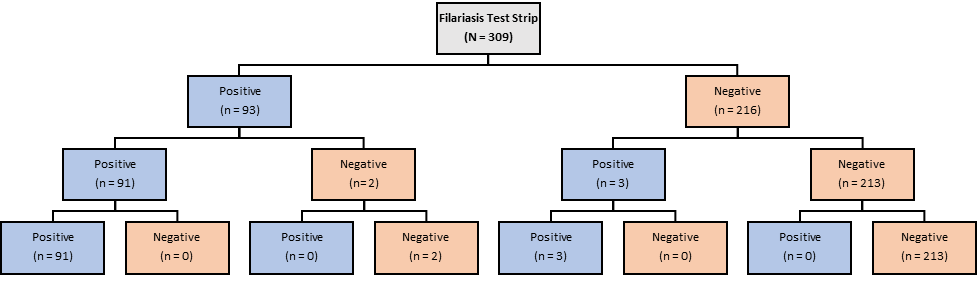

**A**

**10 minutes**

**1 hour**

**Next day**

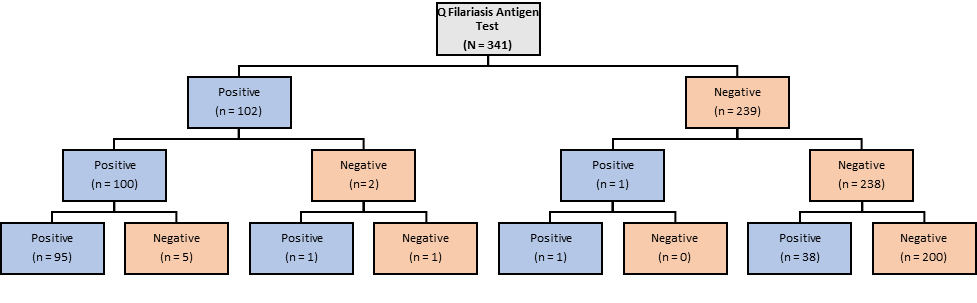

**B**

Filariasis Test Strip (N = 309)

Positive (n = 93)

Positive (n = 91)

Positive (n = 91)

Negative (n = 0)

Negative (n= 2)

Positive (n = 0)

Negative (n = 2)

Negative (n = 216)

Positive (n = 3)

Positive (n = 3)

Negative (n = 0)

Negative (n = 213)

Positive (n = 0)

Negative (n = 213)

Q Filariasis Antigen Test (N = 341)

Positive (n = 102)

Positive (n = 100)

Positive (n = 95)

Negative (n = 5)

Negative (n= 2)

Positive (n = 1)

Negative (n = 1)

Negative (n = 239)

Positive (n = 1)

Positive (n = 1)

Negative (n = 0)

Negative (n = 238)

Positive (n = 38)

Negative (n = 200)
